# Antihypertensive Pharmacotherapy Gaps in Nigeria: A Predictive Machine Learning Analysis of Treatment Uptake Amid Macroeconomic Shock, Using NDHS 2023–24

**DOI:** 10.64898/2026.09.11.26362863

**Authors:** Eloghosa Aisosa Nosa-Ihaza, Emmanuel Chidera Edeh, David Nnamdi Eze, Uyioghosa Nosayise Nosa-Ihaza

## Abstract

**Background:** Hypertension is the most important modifiable cardiovascular risk factor worldwide, and the prevalence is increasing in sub-Saharan Africa. Nigeria’s newly released 2023–24 Demographic and Health Survey (NDHS) provides the first opportunity to explore national-level non-uptake of antihypertensive treatment using a machine-learning cascade framework, although survey fieldwork was conducted amid the unprecedented shock of fuel subsidy removal and currency devaluation in Nigeria.

**Objectives:** To identify correlates of antihypertensive treatment non-uptake (Gap 2) among the diagnosed adults in Nigeria; test if the non-uptake varied based on when the survey was conducted during this macroeconomic shock; and compare four predictive algorithms.

**Methods:** We used the 2023–24 Nigeria Demographic and Health Survey (NDHS) data (2,975 women diagnosed with hypertension and 529 men diagnosed with hypertension aged 15-49) to fit survey-weighted logistic regression models separately by sex and evaluated sex-by-predictor interactions in a single pooled model. We then compared the survey-weighted logistic regression model with the elastic net, random forest, and gradient boosting (XGBoost) models, all properly weighted, using held-out test-set AUC.

**Results:** The timing of fieldwork interviews was a significant risk factor for women (OR=1.14 per month; 95% CI: 1.06–1.22; p<0.001) but not for men, and this sex difference was confirmed in the formal interaction test (p=0.016). This study tested the interaction of sex with each geopolitical zone, with the strongest result being a sex-reversed pattern in zones protective for women, showing these zones were significant risk factors for men (ORs 2.28–5.77); interaction testing (all p≤0.007) confirmed this. No interaction was observed between diabetes and sex (p=0.170); instead, diabetes was protective for women (OR=0.34, p<0.001). For pooled samples, wealth was associated with non-uptake (OR=0.59; p=0.012), and this appeared significant only for women (OR=0.62; p=0.026), in keeping with the richest-vs-poorest estimate in Table 2. All three ML algorithms had similar AUC for women (0.61–0.63) and lower, near-chance performance for men (0.56–0.59), reflecting the much smaller number of diagnosed men.

**Conclusions:** There is a large disparity in the rate of treatment and management of HTN across sexes, geopolitical zones, and macro-economic environments, making a single uniform national intervention model impractical. In contrast, an earlier study in Africa reported that similar algorithmic complexity was useful in predicting cardiovascular risk; this did not happen in the current study. The proposed programs are to build programs by sex and zone, integrate hypertension screenings into chronic disease and/or maternal health trigger points, and better understand the resilience of the pharmacy supply chain.

## 1. Introduction

Hypertension is the most common modifiable risk factor for cardiovascular mortality in the world and not everyone is equally affected by this risk. According to the World Health Organization (WHO, 2023), hundreds of millions to more than a billion adults in the world have high blood pressure, while one-quarter have a good blood pressure level. This burden is also changing: Pooled analyses over the past decades in over 100 countries have shown that NCD prevalence has decreased in many high-income countries and increased in many countries in sub-Saharan Africa and South Asia (NCD-RisC, 2021). They have an increasing burden of hypertension – this is also divergent with Nigeria, the population of Africa’s most populous nation, which exceeded 220 million at the time of this survey (World Bank, 2024, World Development Indicators), as they too have an increasing burden of hypertension, yet the healthcare system is still maturing and under-resourced to detect and treat hypertension at scale.

The steps in a person’s hypertension care pathway—screening, detection, treatment, and control—are commonly called the hypertension care cascade, and the proportion of patients lost along the way is the standard measure of its success. This study is restricted to the second of these gaps: treatment non-uptake among adults with diagnosed hypertension (Gap 2 — the percent of adults with diagnosed hypertension who do not currently take antihypertensive medication). The recently released Nigeria’s Demographic and Health Survey (NDHS) 2023–24, however, does not have a directly measured blood pressure reading; rather, its items related to hypertension are limited to questions related to self-reported screening, self-reported diagnosis, and self-reported treatment (National Population Commission [Nigeria] and ICF, 2024). The scope of this study is therefore limited to Gap 2 alone, a true constraint of available data, rather than one that was imposed as part of the design, as it allows for a policy-relevant analysis of the most actionable of the cascade stages, namely Gap 2, directly actionable by the pharmacy practice and primary care setting.

The access architecture of pharmaceuticals in Nigeria differs markedly from settings previously examined in this literature, making it a key point of differentiation in the chain. Most Nigerians access medicines through a vast, fragmented network of patent medicine vendors and private retail pharmacies, while a relatively small proportion have formal insurance coverage through the National Health Insurance Authority (NHIA) or state-level schemes. Thus, knowing the percentage of diagnosed adults not engaging in consistent therapy after a hypertension diagnosis is directly relevant to the role of community pharmacy, medicine vendor law, and insurance coverage in expanding access to interventions to address this gap.

The third unusual aspect of this survey round provides an opportunity for this study. The fieldwork for NDHS 2023–24 took place during the most severe macroeconomic disruption in Nigeria in a generation, from December 2023 to May 2024 (National Population Commission [Nigeria] and ICF, 2024).The national fuel subsidy removal and shift to a floating exchange rate for the naira in late May 2023, resulted in a dramatic devaluation of the currency, which contributed to the sharp increase in the headline inflation and consequently impacted the cost and availability of imported goods including pharmaceuticals (IMF, 2023). The frequency with which respondents were interviewed in an exact month allows for an opportunity to test whether the likelihood of treatment not being taken by diagnosed Nigerians changed measurably over time while controlling for geographic zone, which is a rare natural-experiment opportunity. This is the first hypertension cascade study we are aware of that has done so.

This gap exists along with two previously identified gaps in the overall literature. Comparative and multi-country analyses of hypertension prevalence and control are not determinant-focused, but rather trend-focused, and often combine multiple countries into country blocs or groups instead of detailing each country’s cascade (NCD-RisC, 2019, 2021). DHS-based cascade studies detail prevalence and treatment gaps within countries but, to our knowledge, have not analyzed fieldwork timing as an exposure. The one work we are aware of that has applied multiple machine-learning algorithms to hypertension cascade outcomes — a similar design with four algorithms applied to Mexico’s ENSANUT survey (Mendoza-Cano et al., 2025) — was applied to a survey conducted in the absence of a similarly dramatic, dateable economic shock.

This study consequently has three objectives. The first objective was to describe the sociodemographic, health system access, and comorbidity correlates of antihypertensive treatment non-uptake separately for women and men, using a survey-weighted logistic regression model. Second, to determine if there were systematic variations in treatment non-uptake based on the timing of the interview in the survey in Nigeria 2023–24, as a direct test of real-time economic shock transmission to chronic-disease medication continuity. Third, to apply a multi-algorithm comparison design, similar to that used in the most comparable previous methodological study, an ENSANUT Mexico-based hypertension cascade study (Mendoza-Cano et al., 2025), to a national context that experienced a concurrent, dateable macroeconomic shock.

## 2. Literature Review

### 2.1 Global and Regional Hypertension Burden

While there has been a slight improvement in the proportion of people with hypertension whose blood pressure is controlled in countries worldwide, most countries have less than 25% (WHO, 2023), and the burden of hypertension is moving from high- and middle-income countries to low-income countries, although a significant proportion of high-income countries experienced substantial reduction (NCD-RisC, 2021). Nigeria is in this swing: The prevalence of adult hypertension, as reported by national and subnational studies, ranges between 25-40%, depending on the setting, sampling frame, and diagnostic criteria used (Ogungbe et al., 2024).

### 2.2 The Hypertension Care Cascade Framework

Geldsetzer suggests that the hypertension care cascade (screening, diagnosis, treatment, control) is the traditional approach to counting where health systems lose patients (Geldsetzer et al. 2018). It is used with nationally representative data in India (Amarchand et al., 2022), with an extended cascade and adherence stages in Sri Lanka (Rannan-Eliya et al., 2025), and in urban-poor communities in Accra, Ghana (Sanuade et al., 2025). These studies offer the cascade as a well-documented, internationally comparable framework with no regard for the timing of fieldwork as an analytic variable, which is explored further in section 2.5.

### 2.3 Cascade Evidence From Nigeria: National and Subnational Studies

A significant amount of country-specific research has used a cascade approach, although, again, most of it is at the sub-national or facility level rather than a nationally representative, individual-level regression estimate such as the one in this study. Across all steps of the pre-intervention cascade in the COMAAND community intervention project (Danladi et al.,2025), dropout percentages at each stage were comparable to those in the current study. This study’s design was sex-stratified, but a similar sex bias was observed in the raw prevalence of diagnosed hypertensives: 34.7% of 811 adults in a single city opportunistic screening sample were aware of their diagnosis, versus 13.8% of all hypertensives (not diagnosed), with a difference of p=0.015 (Amadi et al., 2026). A prior study in Lagos also showed poor uptake of hypertension care after community screening events and suggested a decentralized, pharmacy-based approach to care as a solution (Nelissen et al., 2018)—directly relevant to the concept developed in Section 5.4 on the implications for pharmacy practice. A supply-side study, in terms of the capacity of primary health care centers to provide services, rather than the demand-side determinants of individual patients, is the study of facility readiness, by which the present study is a complement to the literature reviewed, especially the blood pressure study by Orji et al. (2021). A trial protocol for the resulting Hypertension Treatment in Nigeria (HTN) Program, based on the WHO HEARTS package, is described separately (Baldridge et al., 2022). This is an older study (Chijioke et al., 2016) conducted at a facility that had an awareness and control rate significantly higher than the community rates (e.g., 91.9% awareness, as compared to much lower rates in the community), reflective of the well-known selection bias of facility samples towards patients already in care, which this study’s nationally representative DHS design avoids. Community health workers are also proposed to provide hypertension management services in Nigeria (Oseni et al., 2024), a form of outreach service delivery relevant to the male-specific outreach services discussed in Section 5.4. The treatment and control rates that these programmatic initiatives report for their cohorts of patients (described in the text above, for the HTN program, and in Ogungbe et al., 2024, for the National Hypertension Control Initiative, which was piloted in Ogun and Kano states) are not directly comparable to the rates reported in this study, but do provide a useful benchmark for what can be achieved by programmatic care, which in these initiatives was much better equipped than the care available to the general diagnosed population in the study.

### 2.4 Nigeria’s Pharmaceutical Access Architecture: PPMVs and Health Insurance

Nigeria’s medicines-access landscape differs from what is observed in most contexts in the hypertension cascade literature. One source of medicines access that is widely documented is the activities of the patent and proprietary medicine vendors (PPMVs), which are also known as non-pharmacy-trained retail drug vendors, who are regulated by the Pharmacists Council of Nigeria (PCN) (Beyeler et al., 2015; Prach et al., 2015). PPMVs are reported as the initial source of care for 55% of the rural population’s illness episodes among under-5s and 35–55% of the treatment episodes for adults with malaria, nationally, with more than 200,000 outlets (Prach, Treleaven, Isiguzo, & Liu, 2015; Barnes, Chandani, & Feeley, 2008). It is also relevant to understanding this study’s outcome in Gap 2, a largely decentralized and unregulated retail environment, where antihypertensive medication would be accessed by people who have been diagnosed with hypertension but are not accessing formal pharmacy or clinical follow-up, which is not captured by the NDHS and is a specific limitation that should be noted explicitly in Section 5.5.

Formal health insurance coverage in Nigeria remains low compared with many other LMICs. The National Health Insurance Authority (NHIA), which superseded the National Health Insurance Scheme (NHIS) established in 1999, had reported enrolment of some 19.2 million Nigerians in 2024 and 21.7 million in 2025, representing approximately 13% of the Nigerian population (Federal Ministry of health and Social Welfare, 2025), while a peer-reviewed multi-site study found less than 10% of the Nigerian population enrolled in either NHIA or state-supported NHIS as of 2024 (Effiong et al., 2025). Other issues discussed in the Nigerian healthcare policy literature include implementation challenges under the NHIA Act (2022), such as low funding priority, a shortage of healthcare workers, and enforcement of the Act, including forced enrollment (Ipinnimo et al., 2022).

### 2.5 Economic Shocks and Medication Continuity: An Established Literature, Applied Here in a New Way

This study examines a relationship between acute macroeconomic shock and continuity of chronic disease medication that is less studied, but more familiar conceptually. The economic crisis has produced a well documented body of work in Lebanon such as a cross sectional study of 156 Lebanese adults with diabetes or hypertension, which found the economic crisis to have a direct impact on drug shortages, unaffordable healthcare, and disrupted health seeking behavior (Cherfane et al., 2024); a validated scale “Harmful Impact of Medication Shortage” specifically developed to measure this phenomenon (Bou Malhab et al., 2025); and a series of Lebanese studies that examined the general picture of outpatient drug availability and affordability trends through the crisis (Jaber Chehayeb et al., 2023). In the economic crisis in Sri Lanka in 2022, the percentage of people who said that they would change how they used their meds during this time frame ranged between 39–41% of those that were surveyed, with cost being the single most important factor (Jayawardena et al., 2023); and a national US survey following the 2008 financial crisis found substantial cost-related medication non-adherence among chronically ill adults, particularly those who were unemployed or looking for work (Piette et al., 2011). The review of unplanned ARV treatment interruptions that examined acute economic and political shocks to chronic-disease treatment continuity during three southern African crises (in Mozambique in 2008, Zimbabwe’s economic and political collapse, and a public-sector strike in South Africa in 2007) is most relevant for an African context, but it was conducted on HIV, not on hypertension (Veenstra et al., 2010).

This study differs from the above in that those studies use retrospective self-reports of behavior change for a known crisis period, using a survey tailored to that crisis. The study takes a different approach and tests whether a real-time, dateable macroeconomic event can be found as a natural experiment in ordinary cascade data, which was not designed to study the economic shock of 2023-24. As far as we know, no other hypertension-cascade study has been designed and conducted like this one, but that does not necessarily mean so, since the search for this review was not systematic or exhaustive, and merits further investigation if a more similar precedent is found during peer review.

### 2.6 Machine Learning Applications to Hypertension, Including in Nigeria and Africa

A recent systematic review of ML applications to hypertension in Nigeria identified six eligible primary studies published between 2017 and 2025 (Uzoechina et al., 2025). Four were described in enough detail to compare directly: a university workforce study using unsupervised clustering only (n=1,723); a community-based study using an artificial neural network for binary diagnosis (n=211); a clinic-based study comparing random forest, XGBoost, and SVM to predict therapy type rather than diagnosis or treatment uptake (n=303, the richest of the algorithmic comparisons among these six); and a community-based study using CART and ANN for binary systolic blood pressure classification (n=201), which the review flagged as high risk of bias given its implausible 100% accuracy in a dataset of only 22–32 records.

None of these six studies is based on a nationally representative, complex-survey-design dataset with sampling weights, PSUs, or strata, and the richest of the algorithmic comparisons among these studies (Akhaine, three algorithms) does not use logistic regression or elastic net as baseline comparators, and does not examine treatment non-uptake among diagnosed adults (all predict hypertension diagnosis itself, or, in one case, therapy category). The review’s PROBAST risk-of-bias assessment assigns all six studies a moderate to high risk of bias because of small sample sizes, lack of external validation, and—in two instances—near-perfect internal validity results, which in turn indicate severe overfitting. The research-gap and future-directions sections of its own published paper explicitly request “nationally representative data” and “comparative evaluation of ML and classical statistical models,” respectively — both of which this study’s design provides. This study’s diagnosed-women subsample (n=2,975) is larger than all six previous Nigerian counterparts combined.

However, the African and global ML-hypertension literature is more developed than the ML-hypertension literature in Nigeria specifically. A similar study using WHO STEPS survey data from 57 countries and six WHO regions found that the five algorithms (logistic regression, k-NN, random forest, XGBoost and a neural network) clustered closely — a convergence of algorithms seen in the Sub-Saharan Africa subset of the study, for which the four comparable algorithms (Bisong et al., 2024) found a similar convergence of algorithms, where elastic net, random forest and XGBoost performed similarly on Nigerian women. More directly comparable in algorithm choice, a 2026 study published in medRxiv used WHO STEPS data from 60,294 adults from across 12 African countries (Ng’ambi et al., 2026), and the same three algorithms as used here (elastic net, random forest, XGBoost) to predict cardiovascular disease risk, with XGBoost showing the highest discrimination (AUC=0.769) — a direct point of contrast worth engaging with substantively in the Discussion, as that study found tree-based complexity paying off where this study’s results did not. There are also some country-specific works: in Ethiopia (Islam et al., 2023) and a study consisting of three countries in South Asia (Islam et al., 2022). Previous studies have modeled the under-five mortality cascade in Nigeria using machine learning, paving the way for the analytic approach used in this study (Samuel et al., 2024); however, no prior study has modeled the hypertension cascade in Nigeria using machine learning.

### 2.7 Synthesis: The Gaps This Study Addresses

These three gaps have driven this study and are now directly addressed. First, the existing Nigerian literature is limited to facility- or sub-national-level studies of program-retained cohorts of diagnosed hypertensives, rather than a nationally representative estimate for the general population of diagnosed adults (see Section 2.3). Second, although some evidence suggests other shocks cause medication non-continuity (section 2.5), no study has tested this concept as a natural experiment using within-survey medication non-continuity driven by interview timing variation. Third, and now directly demonstrated, in Nigeria, no prior study has combined a nationally representative survey-weighted design with a four-algorithm comparison for treatment non-uptake among diagnosed adults; the present study’s sample of diagnosed women is nearly 75% larger than the largest prior Nigerian ML study on this topic.

## 3. Methodology

### 3.1 Study Design and Data Source

This study employed a cross sectional design and a national representative household survey carried out by National Population Commission (Nigeria) in collaboration with the Federal Ministry of Health and Social Welfare, in partnership with ICF, using Technical assistance from ICF under the DHS Program (National Population Commission [Nigeria] and ICF, 2024). The fieldwork was undertaken from 1st December 2023 to 7th May 2024 and was based on multistage stratified cluster sampling of 1400 primary sampling units (PSUs) and 42,000 households in all 36 states and the Federal Capital Territory (FCT). These are two individual-level files from the survey: the Individual Recode (women’s questionnaire, IR), which has 39,050 women aged 15–49; and the Men’s Recode (MR), with 12,204 men aged 15–59. While a structural feature of the sampling design, and not a limitation of this analysis, the men’s questionnaire was administered in just one in three of the households selected for the women’s questionnaire (National Population Commission [Nigeria] and ICF, 2024), which explains a significant portion of the size difference between the two analytic files.

### 3.2 Study Population

Two analytic subgroups were created, one for each sex, limited to individuals diagnosed with hypertension (as defined in Section 3.3). For women, this represents an analytic sample of 2,975 diagnosed adults from 39,050 women interviewed. For men, two population definitions are used: one including ages 15–49 (n=529), the same age range as the women and the age range used in the published tables for hypertension in the Final Report; a second using the full age range of the males surveyed (15–59, n=754). The 15-49 age group of males is the main sample used for comparison with the published benchmark statistics in the Final Report, and the full-range sample of males is a secondary sample specification to check the sensitivity of the results to this restriction. All analytic samples, including the subsequent train/test partitions in Section 3.6, are summarized in Figure 1.

**Figure 1.**
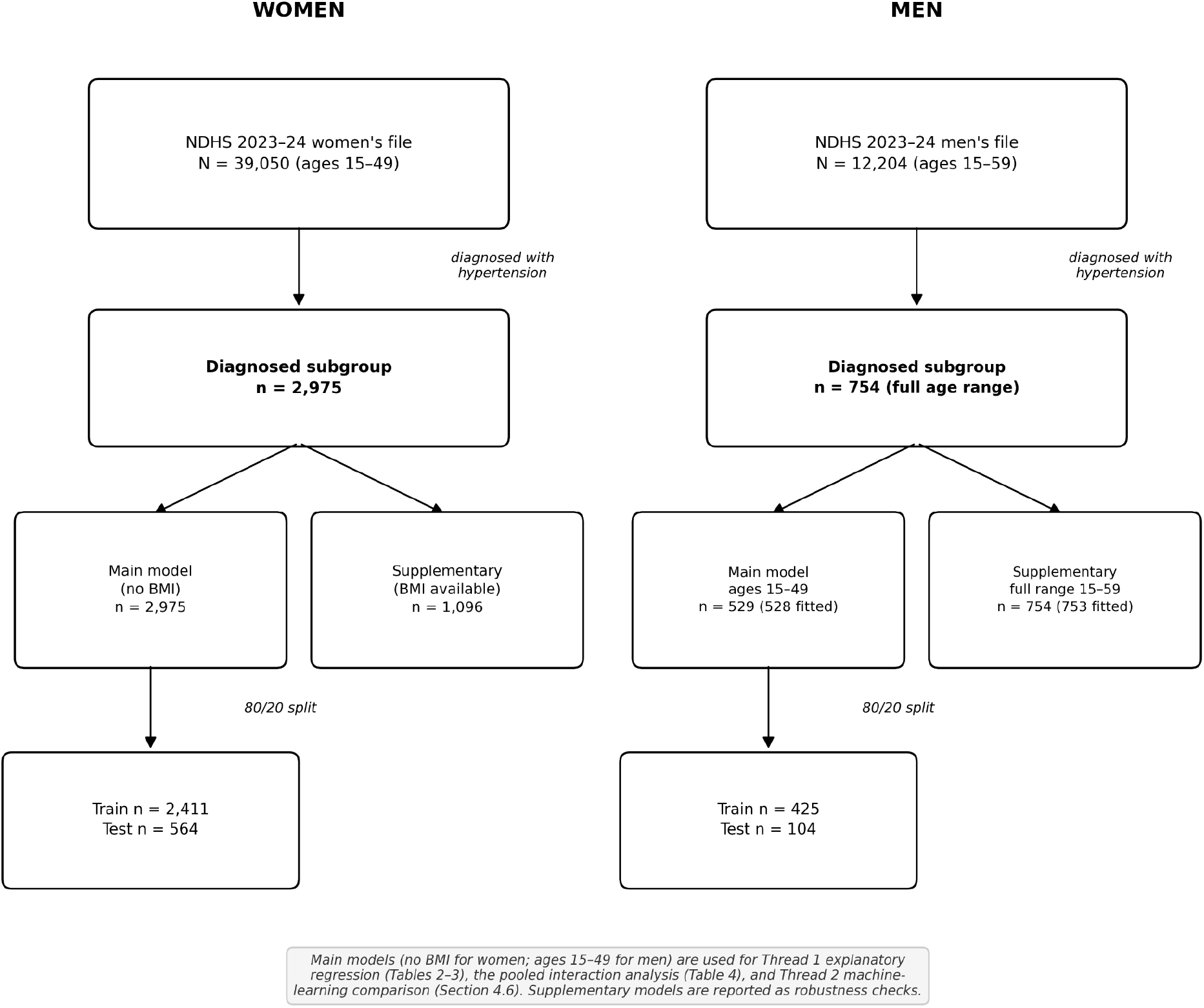
Sample flow diagram.

### 3.3 Outcome Definition

Blood pressure was not directly measured in this survey round, so we do not analyze blood pressure control here. This is not assumed, but three distinct points are highlighted: The DHS-8 Model Biomarker Questionnaire, which structures the content of blood pressure measurements in the DHS surveys generally speaking, does not include a section on blood-pressure measurements for any age group; the complete length of the table of contents of the Final Report does not contain any chapter presenting measured results from the blood pressure measurements (the only device-measured biomarkers reported are anthropometry and haemoglobin in the Nutrition chapter); and the chapter on noncommunicable disease knowledge, attitudes, and behaviour clearly states its intention and content to rely on self-report and not physical measurements.

This study is thus limited to adults who have already been diagnosed with hypertension and do not take their medication. The variables used to create this outcome were the following items from the noncommunicable-disease module of the survey: chd02 (women) and mchd02 (men) (coded “1” if the respondent was ever told by a doctor or health worker that she/he had high blood pressure or hypertension); and chd05 (women) and mchd05 (men) (coded “1” if the respondent was currently taking medication to control her/his blood pressure). We checked the content and categories of both against the country-specific questionnaire reproduced in the appendix to the Final Report, as well as the recode file variable labels. The outcome (Gap 2, treatment non-uptake) is 1 if the treatment flag is negative, and 0 if the treatment flag is positive; the outcome is modeled only within the diagnosed subgroup (the other groups, who have a negative treatment flag, are omitted).

### 3.4 Predictor Variables

The variables used included: age (continuous); educational attainment (no education = reference, primary, secondary, higher); household wealth index quintile (poorest = reference, poorer, middle, richer, richest) using the survey’s own “zone for final report” variable rather than an alternative de jure region-of-residence variable because the former is aligned with the published zone breakdowns in the Final Report and because there is a small “not a de jure resident” category in the latter; religion (Catholic = reference, other Christian, Islam, Traditionalist, Other); ethnicity (Hausa/Fulani = reference, Yoruba, Igbo, Other), which was collapsed from over 300 raw response categories into four analytically tractable groups by directly confirming the underlying numeric codes against the survey’s value labels; health insurance coverage (binary); and a self-reported diabetes comorbidity flag (ever told by a health worker that the respondent has high blood sugar or diabetes).

Body mass index (measured height and weight) was measured in only a subsample of women, and only 1,096 of the 2,975 women diagnosed had this measure, reflecting the design of the anthropometry subsample (rather than item non-response); in addition, a battery of four items that asked about difficulties accessing health care as a “big problem” was measured only in women, as confirmed genuinely absent from the men’s questionnaire.

### 3.5 Fieldwork-Timing Variable

The standard variable ‘date of interview’ coded by the survey in centuries, with a 10-month code, was recoded into a 6-month fieldwork window with a position variable, ranging from 0 (December 2023) to 5 (May 2024). This variable was added not as a nuisance control, but because it was the only one that was observed to have followed exactly the Nigerian economy’s macroeconomic shock of 2023, which was triggered by the removal of fuel subsidies and the floating of the Naira currency, such that it saw a rapid increase in headline inflation through the entire survey period (IMF, 2023). We then cross-tabulated the geopolitical zone with fieldwork month to eliminate potential timing effects from uneven geographic distribution; interview distributions across the fieldwork period were similar across all six geopolitical zones, with none oversurveyed during the survey months.

### 3.6 Statistical Analysis

**Survey-weighted explanatory modeling** We conducted analyses separately for the women’s and men’s analytic files and accounted for normalized individual sampling weights, primary sampling units, and sampling strata in the survey’s complex sampling design. Because some key associations differ meaningfully by sex (as indicated in Section 4), we modeled them separately by sex using survey-weighted logistic regression. For women, we used two specifications: one without BMI (N=2,975, full diagnosed sample) and one with BMI (N=1,096, anthropometry subsample). We used the latter specification because BMI data were available, not by preference. Two specifications were also estimated for men, one excluding the age category ‘singleton’ (n=529, n=528 entered into the fully adjusted model after the automatic exclusion of the single-age category by the estimation procedure, due to perfect prediction), and the other including the full range of ages, 15-59 (n=754, n=753 adjusted). For each model, adjusted odds ratios (AOR), design-based linearized standard errors (dbLS), 95% confidence intervals (CI), and design-based Wald F-test (dbF) are presented.

**Interaction analysis** Several predictors (wealth, diabetes, fieldwork timing, and the geopolitical zone) were statistically significant in the women’s model but not the men’s model; a pooled analysis of the 15-49 women and men who were diagnosed was conducted to test formally whether there were genuine sex differences, rather than relying on the weaker inference of comparing significance across two separately estimated models. The pooled model included sex as a covariate and all four predictors; however, for men, the predictors with divergent patterns were unavailable and were excluded.

**Predictive validation and comparison of algorithms** We created an 80/20 train-test partition within each sex of the sample diagnosed by the main model (women: n=2,975; men: n=529), using a fixed random seed for reproducibility, to compare out-of-sample predictive performance. The four methods of modeling were compared, with survey-weighted logistic regression as the baseline population-representative method and the three machine-learning algorithms (elastic-net-penalized logistic regression, random forest, and gradient boosting (XGBoost)) fit in Python with the scikit-learn package (Pedregosa et al., 2011) and XGBoost (Chen & Guestrin, 2016). All three machine-learning algorithms were fit using the survey sampling weights directly, rather than relying on an unweighted approximation. In addition, we trained each algorithm’s hyperparameters on the training partition only using five-fold cross-validation, optimizing the area under the receiver operating characteristic curve (ROC). Moreover, the hyperparameters of each algorithm were optimized using five-fold cross validation on the training partition only, based on the area under the receiver operating characteristic curve (ROC): elastic net (regularization strength C ∈ {0.01, 0.1, 1, 10}; L1 ratio ∈ {0.1, 0.3, 0.5, 0.7, 0.9}); random forest (number of trees ∈ {200, 500}; maximum depth ∈ {3, 5, 10, unrestricted}); gradient boosting (number of trees ∈ {100, 300}; maximum depth ∈ {3, 5, 10, unrestricted}).; gradient boosting (number of trees ∈ {100, 300}; maximum depth ∈ {3, 5}; learning rate ∈ {0.01, 0.1}). We used AUC as the sole measure of final performance, measured on the held-out test partition. We retrieved variable importance from the random forest and gradient boosting models and used it for qualitative comparison with the logistic regression ORs.

### 3.7 Software

We managed the data, declared the survey design, and ran the logistic regression models (explanatory and interaction) in Stata (version 15). We implemented the machine learning comparison in Python (version 3.9), using scikit-learn for elastic net and random forest modeling and cross-validation, and xgboost for gradient boosting. All machine learning comparisons were carried out in Python (version 3.9), using pandas for data handling; scikit-learn for elastic net and random forest modeling and cross-validation; and xgboost for gradient boosting.

### 3.8 Ethical Considerations

No primary data were collected in this investigation; it is a secondary analysis of anonymous publicly available survey data. All survey participants provided informed consent prior to interviews, and the survey instruments and protocols for NDHS 2023–24 were approved by the ICF Institutional Review Board and the Nigeria National Health Research Ethics Committee (National Population Commission [Nigeria] and ICF, 2024). This study is a secondary analysis of anonymized data gathered with this existing ethical approval and therefore did not require further institutional review.

## 4. Results

### 4.1 Baseline Characteristics

Table 1 presents the results of the comparison between diagnosed adults taking antihypertensive drugs (Gap 2 = 0) and those who weren’t (Gap 2 = 1) separately by sex. All comparisons are design-based estimates with valid F tests for women. A technical limitation that applies to men is not left implicit here: the subpopulation of diagnosed men (n=529) was small, and splitting it out for at least one outcome grouping into strata yielded several strata with only one contributing cluster, rendering it impossible to conduct formal design-based significance tests for each row of men in this table. This does not change the regressions in Tables 3–4, which fit the outcome as a single regression across the design rather than splitting the design into groups first. The weighted percentages and means for men below are valid point estimates; we report no p-values.

**Table 1.**
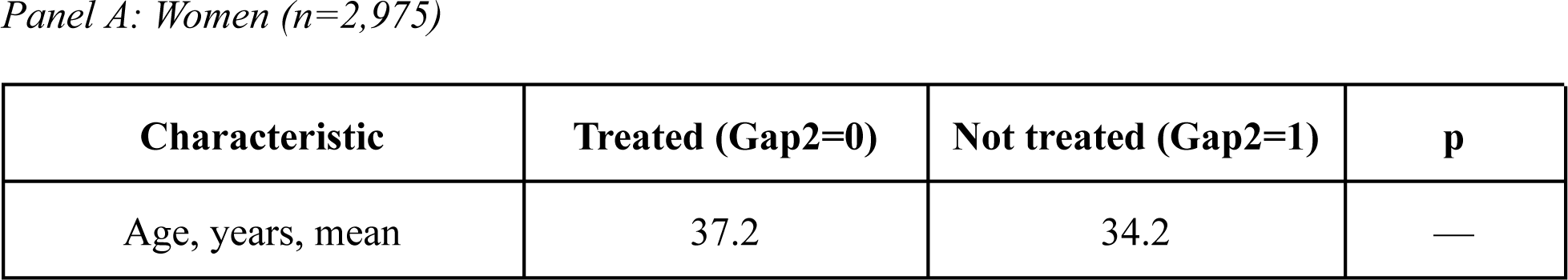
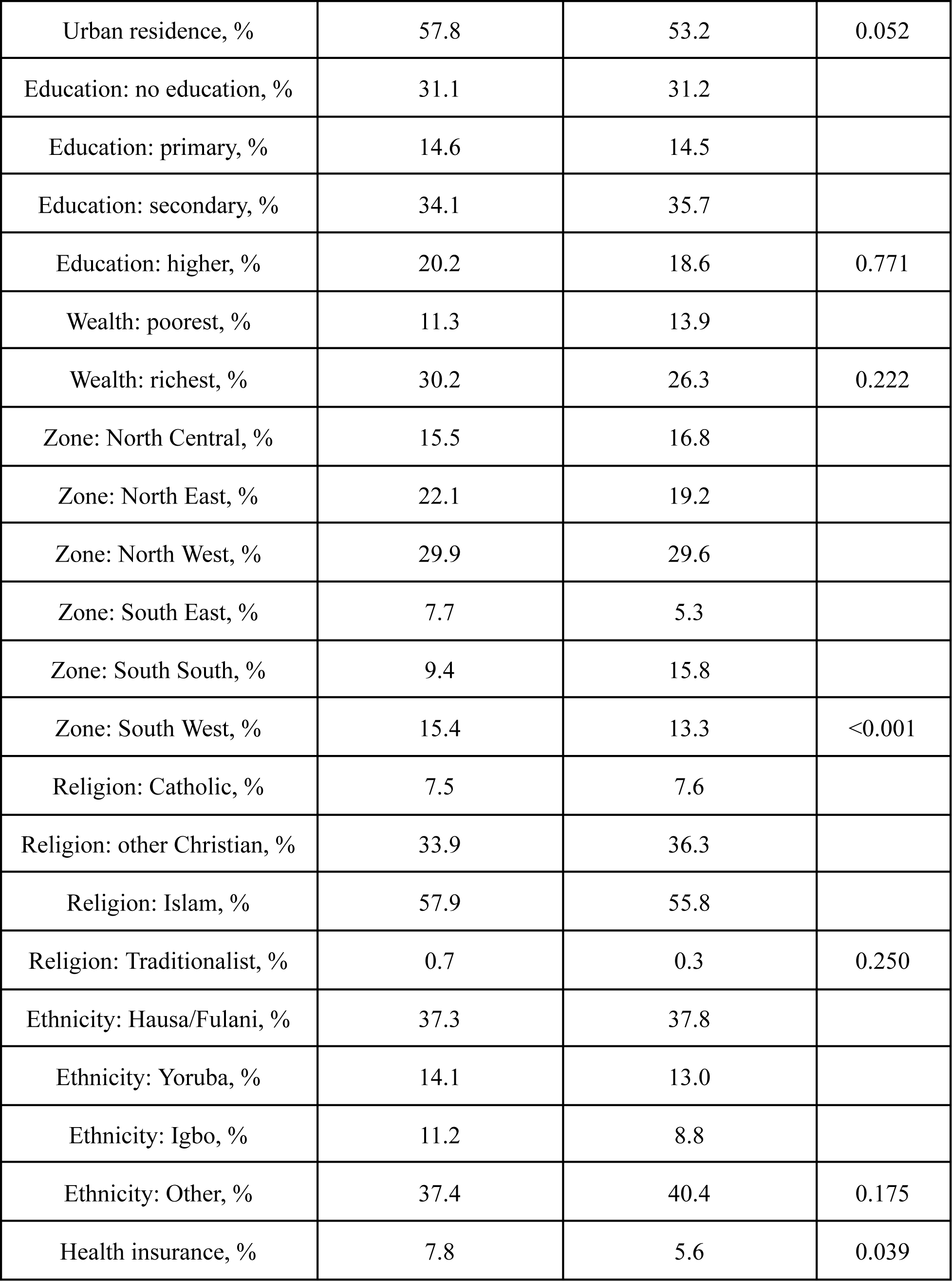
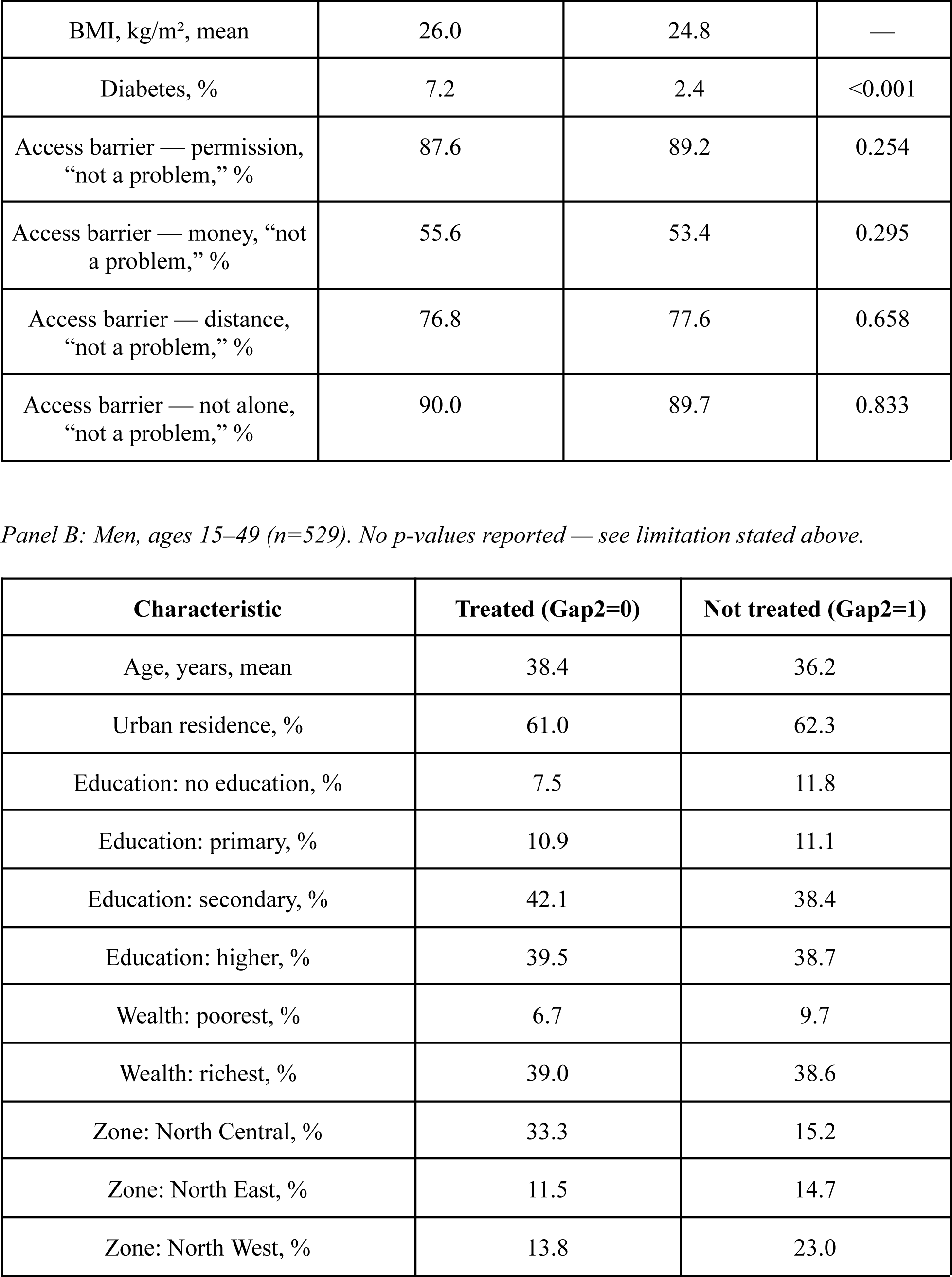
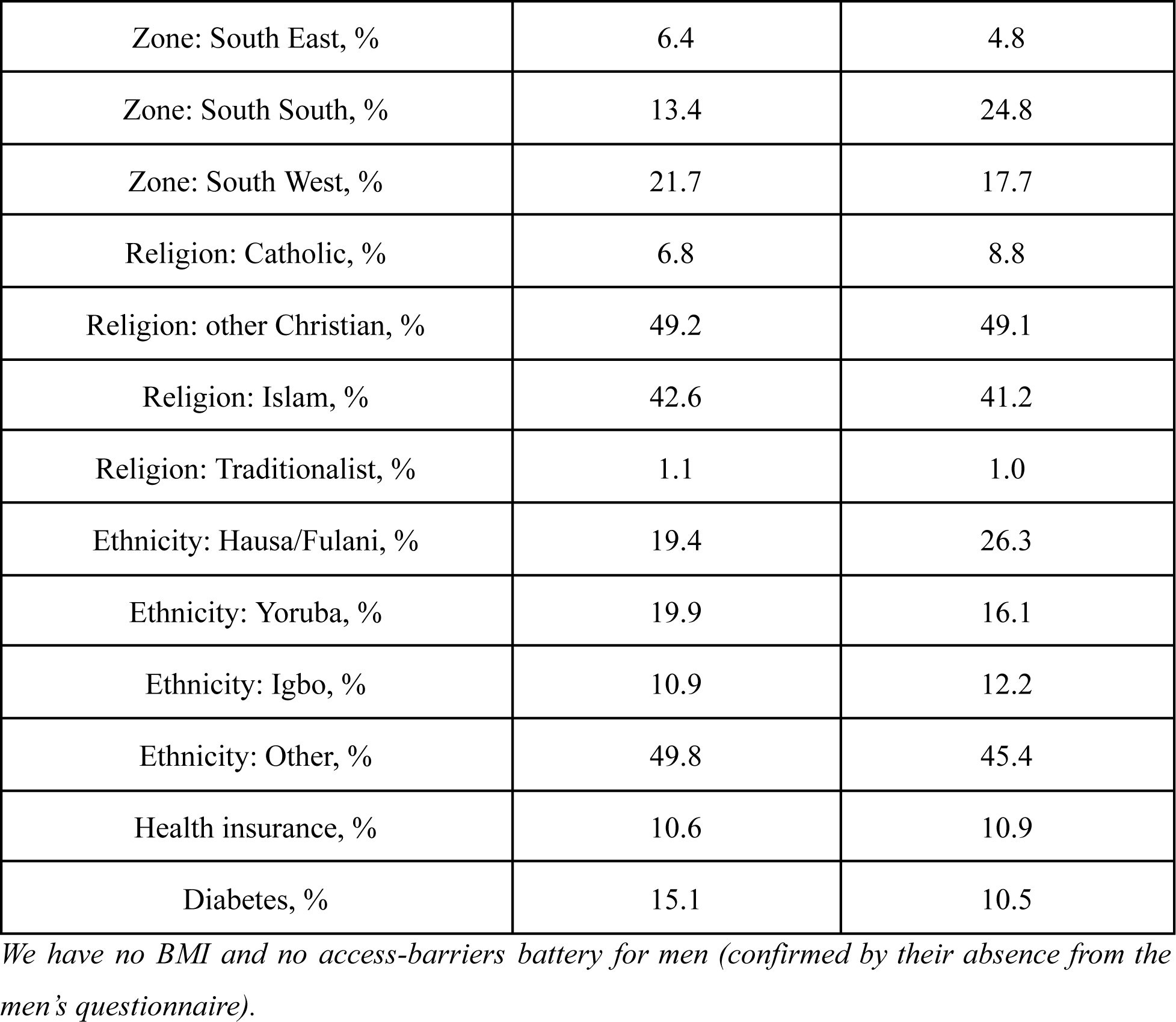
Baseline characteristics of diagnosed adults, by treatment status.

*Panel A: Women (n=2,975)*
| Characteristic | Treated (Gap2=0) | Not treated (Gap2=1) | p |
| --- | --- | --- | --- |
| Age, years, mean | 37.2 | 34.2 | — |
| Urban residence, % | 57.8 | 53.2 | 0.052 |
| Education: no education, % | 31.1 | 31.2 |  |
| Education: primary, % | 14.6 | 14.5 |  |
| Education: secondary, % | 34.1 | 35.7 |  |
| Education: higher, % | 20.2 | 18.6 | 0.771 |
| Wealth: poorest, % | 11.3 | 13.9 |  |
| Wealth: richest, % | 30.2 | 26.3 | 0.222 |
| Zone: North Central, % | 15.5 | 16.8 |  |
| Zone: North East, % | 22.1 | 19.2 |  |
| Zone: North West, % | 29.9 | 29.6 |  |
| Zone: South East, % | 7.7 | 5.3 |  |
| Zone: South South, % | 9.4 | 15.8 |  |
| Zone: South West, % | 15.4 | 13.3 | <0.001 |
| Religion: Catholic, % | 7.5 | 7.6 |  |
| Religion: other Christian, % | 33.9 | 36.3 |  |
| Religion: Islam, % | 57.9 | 55.8 |  |
| Religion: Traditionalist, % | 0.7 | 0.3 | 0.250 |
| Ethnicity: Hausa/Fulani, % | 37.3 | 37.8 |  |
| Ethnicity: Yoruba, % | 14.1 | 13.0 |  |
| Ethnicity: Igbo, % | 11.2 | 8.8 |  |
| Ethnicity: Other, % | 37.4 | 40.4 | 0.175 |
| Health insurance, % | 7.8 | 5.6 | 0.039 |
| BMI, kg/m<sup>2</sup>, mean | 26.0 | 24.8 | — |
| Diabetes, % | 7.2 | 2.4 | <0.001 |
| Access barrier — permission, “not a problem,” % | 87.6 | 89.2 | 0.254 |
| Access barrier — money, “not a problem,” % | 55.6 | 53.4 | 0.295 |
| Access barrier — distance, “not a problem,” % | 76.8 | 77.6 | 0.658 |
| Access barrier — not alone, “not a problem,” % | 90.0 | 89.7 | 0.833 |

*Panel B: Men, ages 15–49 (n=529). No p-values reported — see limitation stated above.*
| **Characteristic** | **Treated (Gap2=0)** | **Not treated (Gap2=1)** |
| --- | --- | --- |
| Age, years, mean | 38.4 | 36.2 |
| Urban residence, % | 61.0 | 62.3 |
| Education: no education, % | 7.5 | 11.8 |
| Education: primary, % | 10.9 | 11.1 |
| Education: secondary, % | 42.1 | 38.4 |
| Education: higher, % | 39.5 | 38.7 |
| Wealth: poorest, % | 6.7 | 9.7 |
| Wealth: richest, % | 39.0 | 38.6 |
| Zone: North Central, % | 33.3 | 15.2 |
| Zone: North East, % | 11.5 | 14.7 |
| Zone: North West, % | 13.8 | 23.0 |
| Zone: South East, % | 6.4 | 4.8 |
| Zone: South South, % | 13.4 | 24.8 |
| Zone: South West, % | 21.7 | 17.7 |
| Religion: Catholic, % | 6.8 | 8.8 |
| Religion: other Christian, % | 49.2 | 49.1 |
| Religion: Islam, % | 42.6 | 41.2 |
| Religion: Traditionalist, % | 1.1 | 1.0 |
| Ethnicity: Hausa/Fulani, % | 19.4 | 26.3 |
| Ethnicity: Yoruba, % | 19.9 | 16.1 |
| Ethnicity: Igbo, % | 10.9 | 12.2 |
| Ethnicity: Other, % | 49.8 | 45.4 |
| Health insurance, % | 10.6 | 10.9 |
| Diabetes, % | 15.1 | 10.5 |

*We have no BMI and no access-barriers battery for men (confirmed by their absence from the men's questionnaire).*

### 4.2 Analytic Samples and Validation Against Published Benchmarks

Of the 39,050 women aged 15–49, 2,975 (7.62% unweighted, 7.97% weighted) indicated that they had been diagnosed with hypertension in the past. This is similar to the published estimate of 8% (National Population Commission [Nigeria] and ICF, 2024). In this diagnosed population, 46.36% (weighted) were not taking any antihypertensive treatment (the report suggested around 46% were taking treatment, which would imply 46% were not). 754 (6.18% unweighted) of the 12,204 men aged 15 to 59 said they had a diagnosis; in the age range used in the report’s own benchmark table (15 to 49), 529 diagnosed men were found. Weighted treatment prevalence was 50.8% (95% CI: 45.9–55.7%) in this age-defined population, similar to the report’s 51% for males. The result variables in this analysis are valid because the consistency between the independently derived weighted estimates and the survey’s published statistics supports this.

### 4.3 Survey-Weighted Logistic Regression: Women

Table 2 shows the main model (without body mass index; n = 2,975 women diagnosed). Older age was protective against non-uptake (OR=0.96, 95% CI 0.94–0.97, p<0.001). High household wealth (top quintile (richest) compared with the lowest quintile (poorest) was protective (OR=0.62, 95% CI 0.41–0.94, p=0.026). Diabetes comorbidity was strongly protective (OR=0.34, 95% CI 0.21–0.54, p<0.001). Non-uptake was significantly higher in South South (OR=1.62, p=0.006) and less significantly for two zones (North East: OR=0.60, p=0.002 and North West: OR=0.65, p=0.020) than the North Central reference zone. The timing of fieldwork was important and substantial: every extra month spent in the fieldwork period raised the odds of non-uptake by 14% (OR=1.14, 95% CI 1.06-1.22, p<0.001); across the full December-April fieldwork period (excluding the very small subsample of May data), the odds of non-uptake approximately doubled. Non-uptake was lower for traditionalist religion (OR=0.32, p=0.004); however, this is based on a small subset of women (193 of the full sample), and should be interpreted accordingly. No significant predictors included education, urban/rural residence, ethnicity, or health insurance.

**Table 2.** Survey-weighted logistic regression, women, main model (n=2,975)

| Predictor | OR | 95% CI | p |
| --- | --- | --- | --- |
| Fieldwork month (0–5) | 1.14 | 1.06–1.22 | <0.001 |
| Age (years) | 0.96 | 0.94–0.97 | <0.001 |
| Education: primary | 0.99 | 0.71–1.37 | 0.947 |
| Education: secondary | 0.94 | 0.69–1.28 | 0.685 |
| Education: higher | 1.03 | 0.72–1.47 | 0.885 |
| Wealth: poorer | 0.79 | 0.54–1.17 | 0.242 |
| Wealth: middle | 0.82 | 0.58–1.16 | 0.262 |
| Wealth: richer | 0.71 | 0.48–1.05 | 0.090 |
| Wealth: richest | 0.62 | 0.41–0.94 | 0.026 |
| Rural | 1.13 | 0.89–1.43 | 0.309 |
| Zone: North East | 0.60 | 0.43–0.82 | 0.002 |
| Zone: North West | 0.65 | 0.46–0.93 | 0.020 |
| Zone: South East | 0.65 | 0.36–1.16 | 0.145 |
| Zone: South South | 1.62 | 1.15–2.27 | 0.006 |
| Zone: South West | 0.78 | 0.53–1.13 | 0.187 |
| Religion: other Christian | 1.03 | 0.75–1.41 | 0.854 |
| Religion: Islam | 0.88 | 0.59–1.31 | 0.520 |
| Religion: Traditionalist | 0.32 | 0.14–0.69 | 0.004 |
| Ethnicity: Yoruba | 1.01 | 0.64–1.59 | 0.979 |
| Ethnicity: Igbo | 0.84 | 0.46–1.53 | 0.565 |
| Ethnicity: Other | 0.82 | 0.59–1.14 | 0.236 |
| Health insurance | 0.86 | 0.61–1.23 | 0.410 |
| Diabetes | 0.34 | 0.21–0.54 | <0.001 |
| Access barrier — permission not a problem | 1.32 | 0.95–1.84 | 0.093 |
| Access barrier — money not a problem | 0.84 | 0.69–1.01 | 0.064 |
| Access barrier — distance not a problem | 1.13 | 0.88–1.46 | 0.345 |
| Access barrier — not alone, not a problem | 0.85 | 0.62–1.18 | 0.332 |

*Reference groups: No education, Poorest wealth quintile, Urban, North Central zone, Catholic religion, Hausa/Fulani ethnicity, No health insurance, No diabetes, Each access barrier reported as "a big problem."*

The supplementary model (n=1,096, restricted to the anthropometry subsample with BMI included as an additional explanatory variable) had a similar overall structure, with fieldwork timing as a significant risk factor (OR=1.13, p=0.043), diabetes as a strong protective factor (OR=0.33, p=0.001), and South South as a significant risk factor (OR=2.46, p=0.001). Body mass index itself was borderline protective (OR=0.97, 95% CI 0.95–1.00, p=0.060). In this smaller sample, the richest-wealth category was not significant in this supplementary model (OR=0.69, p=0.305), which is not unusual when sample size is reduced by about two-thirds, but it is at the lower end of the spectrum.

### 4.4 Survey-Weighted Logistic Regression: Men

Table 3 presents the principal model (n=529; n=528 in the fitted model after automatic exclusion of one case in a single-age category due to perfect prediction, as noted in Section 3.6). Age was protective, consistent with women (OR=0.95, 95% CI 0.93–0.98, p<0.001). Wealth group was not significant. In this latter analysis, the most striking results were for geopolitical zone, which was strongly associated with non-uptake among men: North East (OR=2.28, p=0.039), North West (OR=3.92, p=0.002), and South South (OR=5.77, p<0.001) — the reverse of the pattern seen among women, where these same zones were protective or non-significant (Table 2). The timing of the fieldwork, education, ethnicity, and health insurance were not statistically significant factors.

**Table 3.** Survey-weighted logistic regression, men, main model, ages 15–49 (n=528 fitted)

| Predictor | OR | 95% CI | p |
| --- | --- | --- | --- |
| Fieldwork month<br>(0–5) | 0.94 | 0.81–1.10 | 0.428 |
| Age (years) | 0.95 | 0.93–0.98 | <0.001 |
| Education: primary | 0.76 | 0.28–2.06 | 0.589 |
| Education: secondary | 0.80 | 0.33–1.92 | 0.614 |
| Education: higher | 0.90 | 0.35–2.30 | 0.826 |
| Wealth: poorer | 1.01 | 0.34–3.01 | 0.991 |
| Wealth: middle | 0.73 | 0.23–2.33 | 0.592 |
| Wealth: richer | 0.44 | 0.13–1.46 | 0.178 |
| Wealth: richest | 0.58 | 0.16–2.06 | 0.396 |
| Rural | 0.92 | 0.52–1.61 | 0.765 |
| Zone: North East | 2.28 | 1.04–4.98 | 0.039 |
| Zone: North West | 3.92 | 1.64–9.40 | 0.002 |
| Zone: South East | 1.26 | 0.38–4.24 | 0.703 |
| Zone: South South | 5.77 | 2.79–11.92 | <0.001 |
| Zone: South West | 1.74 | 0.82–3.69 | 0.151 |
| Religion: other<br>Christian | 0.78 | 0.33–1.87 | 0.579 |
| Religion: Islam | 0.72 | 0.27–1.93 | 0.508 |
| Religion:<br>Traditionalist | 0.96 | 0.14–6.81 | 0.970 |
| Ethnicity: Yoruba | 1.39 | 0.46–4.24 | 0.558 |
| Ethnicity: Igbo | 1.50 | 0.37–6.02 | 0.566 |
| Ethnicity: Other | 0.88 | 0.36–2.14 | 0.778 |
| Health insurance | 1.29 | 0.60–2.78 | 0.508 |
| Diabetes | 0.60 | 0.31–1.18 | 0.138 |

The supplementary model was fitted with the full age range (15–59 years) and n=753; this showed a consistent pattern where age continued to be protective (OR=0.95 p<0.001) with all three zone effects again being significant risk factors in similar size (North East OR=1.96 p=0.049; North West OR=3.64 p=0.001; South South OR=4.02 p<0.001), suggesting that the age restriction used in the main model does not influence this finding.

### 4.5 Interaction Analysis: Formally Testing Sex Differences

The data in Sections 4.3 & 4.4 indicate that wealth, diabetes, and timing of fieldwork matter for women but not men, and that the effects of zone are opposite for the two sexes. Table 4 reports the results of formal tests of these patterns, in which diagnosed women (n=2,975) and diagnosed men (n=529) were pooled into a single model (n=3,504; n=3,503 fitted, after excluding the same single case dropped in Section 4.4), and sex-by-predictor interaction terms were added. This formal test does not consistently confirm the pattern indicated by the separate tests, nor are the two tests the same kind of evidence.

| **Interaction** | **OR** | **95% CI** | **p** | **Conclusion** |
| --- | --- | --- | --- | --- |
| Wealth (richest)<br>× sex | 1.20 | 0.43–3.36 | 0.730 | Not supported |
| Wealth (richer) ×<br>sex | 0.72 | 0.25–2.07 | 0.536 | Not supported |
| Wealth (middle)<br>× sex | 0.94 | 0.32–2.70 | 0.902 | Not supported |
| Wealth (poorer)<br>× sex | 1.26 | 0.40–3.95 | 0.689 | Not supported |
| Diabetes × sex | 1.77 | 0.78–3.98 | 0.170 | Suggestive, not<br>confirmed |
| Fieldwork<br>month × sex | 0.82 | 0.69–0.96 | 0.016 | **Confirmed** |
| Zone (North<br>East) × sex | 3.73 | 1.70–8.18 | 0.001 | **Confirmed** |
| Zone (North<br>West) × sex | 5.87 | 2.68–12.85 | <0.001 | **Confirmed** |
| Zone (South<br>East) × sex | 3.00 | 1.34–6.72 | 0.007 | **Confirmed** |
| Zone (South South) × sex | 3.47 | 1.76–6.83 | <0.001 | **Confirmed** |
| Zone (South West) × sex | 2.58 | 1.30–5.13 | 0.007 | **Confirmed** |

No significant interactions emerged between sex and wealth (all p≥0.536). Wealth was also significant in the pooled model (OR=0.59, 95% CI 0.39–0.89; p=0.012), and its association with non-uptake was similar to that in the women-only model, with a similar effect for both sexes. The diabetes-by-sex interaction (OR=1.77) was also directionally consistent with a weaker protective effect among males but was not significant (p=0.170), which may reflect limited power due to the small number of diagnosed, hypertensive males. By contrast, the fieldwork timing interaction was significant (p=0.016), as were all five zone-by-sex interactions (p≤0.007), indicating that these interactions and the zone reversal reflect real sex differences and are not artifacts of comparing two models with different estimation methods. Figure 2 shows these sex-stratified estimates as a forest plot, contrasting the zone reversal and fieldwork-timing divergence — both confirmed sex differences — with wealth and diabetes, which showed more similar effects across sexes.

**Figure 2.**
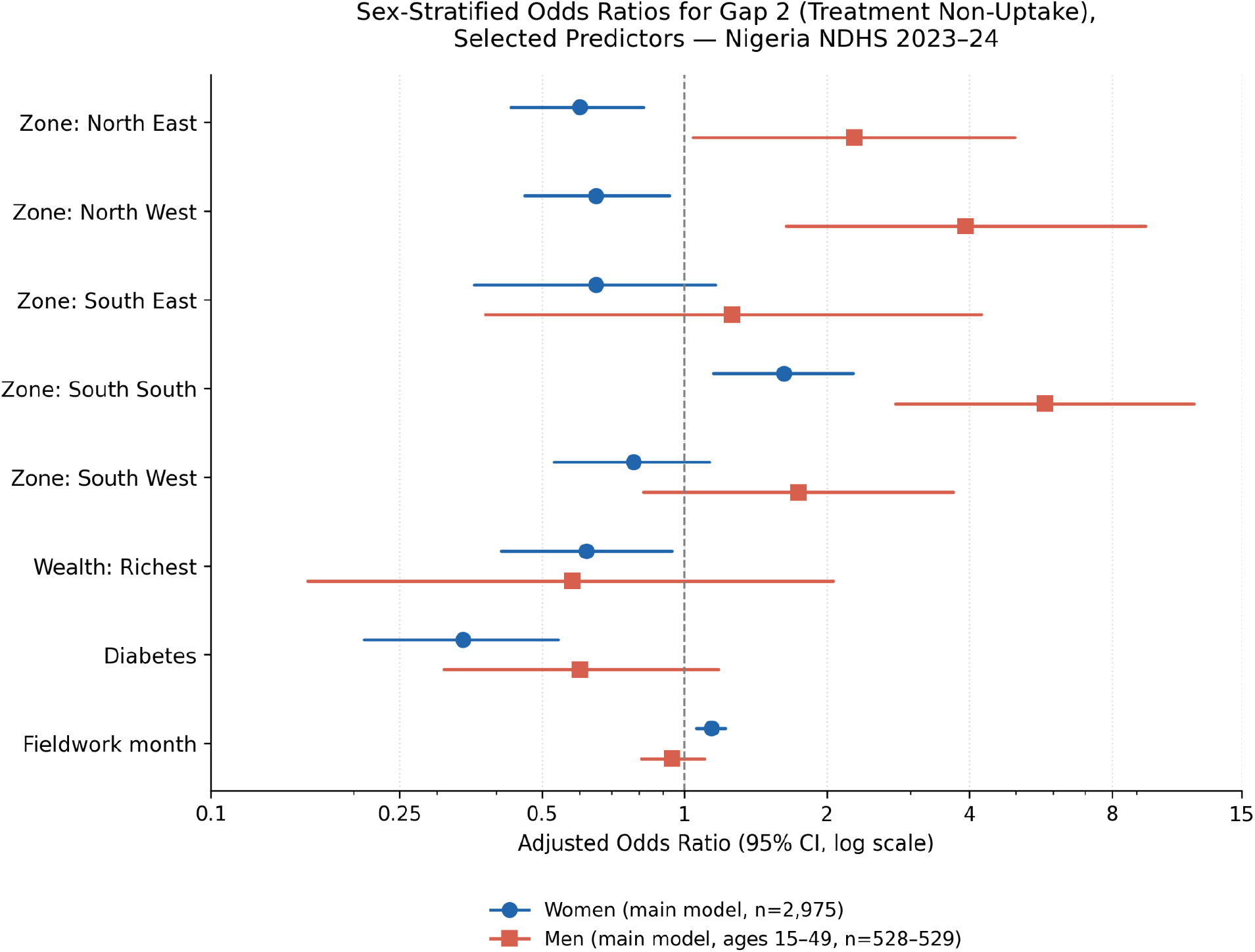
Forest plot of sex-divergent findings.

### 4.6 Predictive Validation: Algorithm Comparison

Figure 3 presents the AUC scores of all three machine-learning algorithms for the held-out test set per sex. For women, the AUC values were very close (elastic net: 0.608; random forest: 0.631; XGBoost: 0.620; all within 0.02 of each other), so no single algorithm clearly outperformed the others. For men (train n=425, test n=104), all three algorithms had lower AUCs and were close to chance (elastic net 0.563, random forest 0.593, XGBoost 0.559) and may not be considered reliable estimates, given the small test-set size.

**Figure 3.**
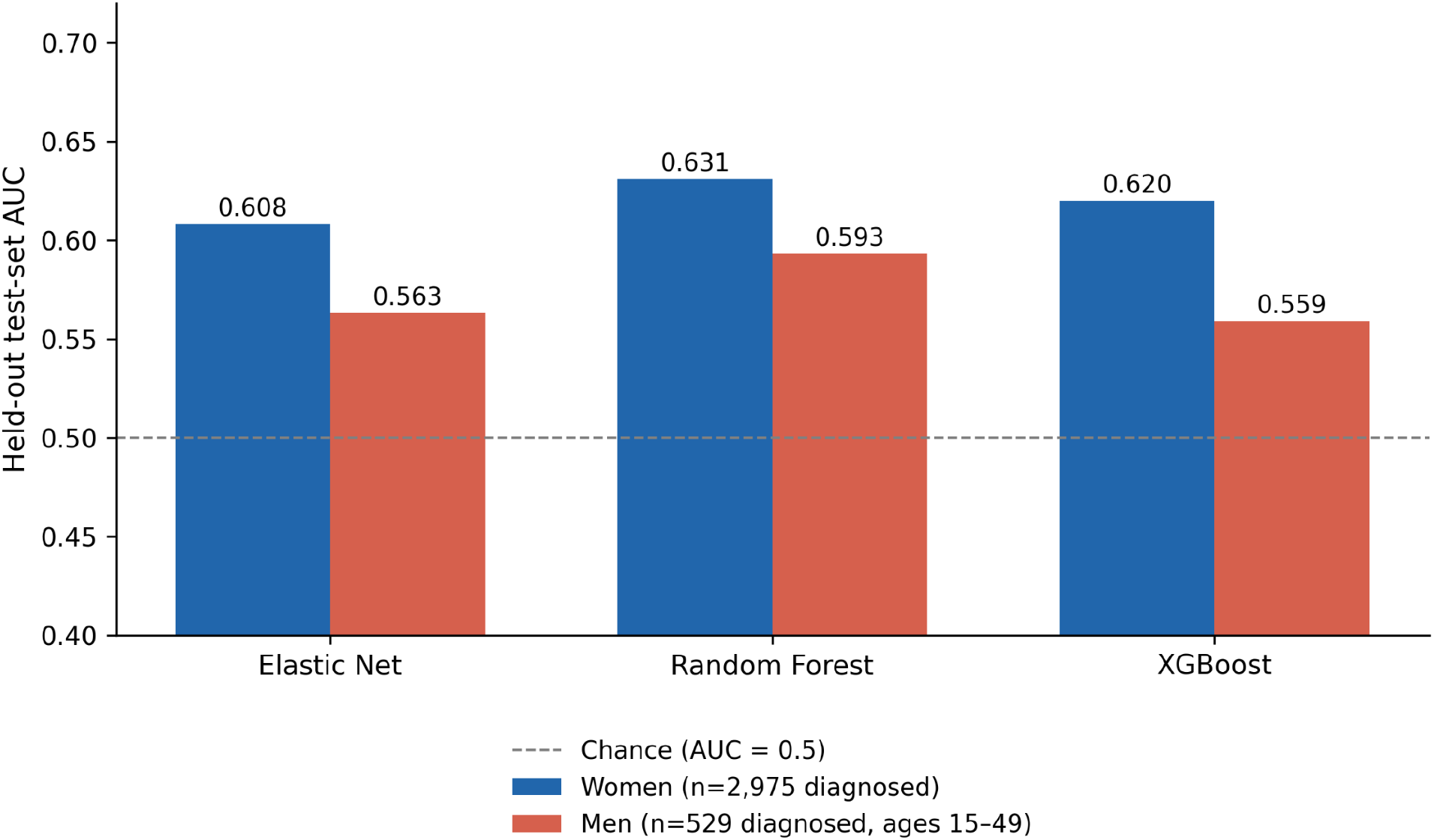
AUC comparison by algorithm and sex.

Variable importance was generally consistent between the two algorithms for women: age, diabetes, fieldwork timing, and the South South zone effect were among the most important variables in both the random forest and the XGBoost results, confirming the logistic regression results in Section 4.3. In the case of men, the two algorithms differed significantly: for random forest, the most important variables were age (importance 0.29) and fieldwork timing (0.13), whereas for XGBoost the most important variables were zone, religion, and wealth, with age close to the bottom (importance 0.05). This difference between two tree-based methods on the same data set is informative in itself: it suggests that the male models are not converging on a clear signal but are instead picking up on different parts of a weak, noisy signal, which is in line with the small sample size of the males (see Section 5).

## 5. Discussion

### 5.1 Principal Findings

The aim of this study was to determine the group of diagnosed hypertensives among Nigerian adults and whether this group is responsive to the acute economic shock at the time of the study. There are four noteworthy findings. First, treatment non-uptake increased measurably as the currency and subsidy crisis continued in real time in Nigeria, and we are not aware of any previous hypertension-cascade study equipped to detect this. Second, we found no timing effect among males, which is suggested by comparing separate models and formally confirmed. Third, the sex-reversed pattern is so strong that it is the largest association across the entire analysis: zones protective for women are risk factors for men, and South South is the only zone harmful for both. Fourth, at the individual level, the three machine-learning algorithms performed comparably to each other in predicting women’s risk, converging on a modest level of discrimination (AUC 0.61–0.63).

### 5.2 Interpreting the Sex-Differentiated Findings

These findings are subject to several important limitations in the interpretation and application.

The zone reversal is the most interesting finding of the paper and, indeed, is so marked that it must be taken with a grain of salt. With the information at hand, this study cannot resolve the various competing explanations. One possible explanation is that women in northern zones have more routine contact with the health infrastructure, such as maternal and reproductive health services as opposed to men, and that such services as antenatal care, family planning, and child immunization provide an incidental pathway to care for women for NCDs that is not available for men in the same zones, so integrating screening for hypertension into existing maternal-and-child-health contacts would help women further but do nothing for men. Another is that patterns of insecurity, internal displacement, or health-worker distribution affect men and women differently in their utilization of care-seeking, a pattern documented elsewhere in Nigeria’s health-access literature but not yet demonstrated for adult hypertension treatment specifically. The finding is consistent with both explanations, and neither of them is supported by it; further work, preferably a qualitative/mixed methods study within the areas of impact, is required before a mechanism can be confidently asserted.

Ideally, the fieldwork-timing finding can be considered in the light of an external shock to the economy, dated and documented (Section 3.5), but the same caveat applies to the lack of the finding among males. The potential is that during acute economic stress, access to antihypertensive drugs is more flexible for women than men, as they might rely more on the informal retail market for drugs, which was dominated by the PPMV sector, than men; or that intrahousehold decisions about who gets what during economic stress may have a greater impact on women’s ability to continue with their antihypertensive treatment than on men’s. The data from this study do not allow a distinction between these accounts, and the sex-differentiated finding should be interpreted as a convincing empirical pattern that requires explanation but does not provide sufficient support for any particular explanation already available.

### 5.3 Comparison with the Broader Machine Learning Literature

A 2026 study was identified in Section 2.6, which showed that the same set of three algorithms (elastic net, random forest, XGBoost) used here, and analyzed cardiovascular disease risk across 12 African countries with data from the WHO STEPS survey, had materially better discrimination for XGBoost than for simpler approaches (AUC=0.769). This differs from this study’s results, where all three algorithms were within 0.02 AUC of each other for women. However, three possible, non-mutually exclusive reasons explain why the two datasets might differ. First, that study’s outcome (cardiovascular disease risk) is a composite made up of multiple continuous clinical measures (blood pressure, cholesterol, glucose, anthropometry) with well-documented non-linear and threshold relations with risk that are appropriate for the tree-based methods that can be used to exploit; this study’s outcome, by contrast, is a binary behavioral one (medication continuation) predicted largely from categorical sociodemographic variables, for which a correctly specified logistic model may capture most exploitable structure. Second, the three subgroups diagnosed in this study are almost certainly much smaller than the pooled 12-country sample in that study; and tree-based methods generally need more data to outperform linear methods — as was the case in this study for the men’s model, with random forest and XGBoost very different from each other, which is more a sign of small-sample noise-fitting than signal discovery. Third, predictor richness varies: the study’s Gap 2 model has less predictor richness than STEPS surveys, which typically include continuous clinical biomarkers (Section 5.5). These are presented as possible explanations – not proven – and the differences between the two studies are themselves a good indicator that the relative importance of algorithmic complexity for prediction in the context of hypertension is not a fixed methodological fact, but is also outcome- and context-dependent.

### 5.4 Policy and Pharmacy Practice Implications

Treatment discontinuity is a risk factor that can be modified by macroeconomic shock exposure, and a policy lever is pharmaceutical supply-chain policy. The women’s fieldwork timing finding suggests that treatment continuity for hypertension in Nigeria is not insulated from shocks to currency and import prices, likely mediated in part by the retail channel through which most diagnosed patients access treatment (Section 2.4): PPMV. This is a case for medicine supply to be a dedicated concern in future planning for macroeconomic reform, such as excusing essential NCD medicines from tariff or exchange-rate pass-through mechanisms, or speeding up local manufacturing capacity for antihypertensive active ingredients and finished products so as to limit the import-price exposure related to NCD medicines as identified in section 2.5.

The zone by-sex reversal case supports the rejection of one uniform national hypertension program and the targeting of sex in existing NCD programs within Nigeria. This study’s results indicate that scale-up sequencing and outreach design should vary not only by zone but also by sex within the zone, with programs like the National Hypertension Control Initiative (NHCI) and the WHO HEARTS-based Hypertension Treatment in Nigeria (HTN) program organized around geographic scale-up. Specifically, this might involve, in the areas where hypertension uptake among women is low, engaging women in hypertension screening activities at existing MCH contact points, and in areas where men’s uptake is low, reaching men in a new way, such as through workplace screening or through community health workers, who were included in the work of other health promotion initiatives in Nigeria’s own literature on NCDs (Section 2.3).

The benefits of diabetes comorbidity as a treatment uptake facilitator among women pave the way for integrated, opportunistic NCD screening, not disease-specific screening programs. Whereas the converse-testing directly and explicitly – opportunistic screening for hypertension in diabetes, HIV, and antenatal care contact points – may deliver a substantial portion of the Gap 2, at a relatively low marginal cost, leveraging existing infrastructure rather than using new contact points to deliver hypertension services to people who would otherwise not know to seek them out. This is not the case for men in this study’s data and further underlines the need to have a different approach to addressing the needs of men and women.

This is likely not directly tested by this study’s data, but it is strongly suggested: community pharmacy and PPMV engagement. Although NDHS does not record it, Nigeria’s literature on medicines access (Section 2.4) clearly shows that most Nigerians obtain (or do not obtain) their medicines from PPMVs and private retail pharmacies. This study frames the intervention within the cascade, appealing to the need to structure antihypertensive dispensing practice, with PPMVs trained in basic antihypertensive dispensing practice, adherence, counseling, and referral, so the intervention does not require new infrastructure and is scalable.

### 5.5 Limitations

A particular constraint is the size of the male analytical sample. Two structural features of the survey are responsible. First, the men’s questionnaire was only administered in one-third of the households sampled for the women’s questionnaire (National Population Commission [Nigeria] and ICF, 2024), before any attrition due to hypertension; a total of 12,204 men were interviewed compared to 39,050 women. Second, the prevalence of diagnosed hypertension is lower in this sample of men than women (5.87% weighted vs. 7.97% weighted); as elsewhere in this survey and elsewhere in the hypertension literature from Nigeria, men are less likely to have contact with health facilities than women, and less likely to have had a blood pressure screening. The resulting analytic sample of 529 diagnosed men aged 15–49 (754 in the full 15–59 age range) is small, which limits the precision of the male-specific estimates in Tables 3 and 4. The lack of evidence of effect for diabetes and fieldwork timing is a realistic limitation of one survey round and not a design failure: the results are more credibly interpreted as inconclusive than as evidence of no effect (Section 4.5 shows that there is a genuine difference by sex in fieldwork timing, and this difference is suggestive but not confirmed for diabetes). Wealth is another case: although the interaction test yielded no evidence of a difference in how this variable operates by sex, the absence of significance among men should not be interpreted as a power-limited null hypothesis, but as consistent with a single pooled effect on both sexes. The high odds ratios among men (which were also confirmed to be significantly different by that same interaction test among men and women for zone alone) still suggest that the male analysis had sufficient power to detect an association of that magnitude, but that may not be the case for smaller associations that would occur in a larger sample.

In addition, there are some additional constraints. This study cannot speak to treatment quality among those who are medicated; it can only speak to treatment initiation, because it did not measure blood pressure in this survey round (Section 3.3). As in all DHS-based cascade research, participants self-report diagnosis and treatment status, which are subject to recall and social-desirability bias. The cross-sectional design permits association but not causal inference, including for the fieldwork-timing finding, which — while a genuine natural-experiment opportunity relative to prior retrospective-crisis-survey designs (section 2.5) — remains an association robust to one specific alternative explanation (geographic rollout, ruled out in Section 3.5) rather than a design that rules out every possible confound. Predictor richness for men is lower than for women, since body mass index and the health-care-access-barriers battery are both absent from the men’s questionnaire (Section 3.4), which may partly explain the weaker, less stable predictive performance found for men in Section 4.6. Lastly, the ethnicity variable was reduced from more than three hundred categories to four broad categories (Section 3.4), which was needed for stability of estimation, but at the same time obscured the heterogeneity in the large residual category of “Other,” which would have been better resolved with a larger sample size.

### 5.6 Conclusion

The problem of hypertension pharmacotherapy gap in Nigeria is not a single one, but a multiplicity of problems in space, gender and time. A woman in the North West is not as likely as a man to be missed from treatment, and the same national economic shock is not equally likely to have an impact on both men’s and women’s treatment continuity. These findings contradict the notion of a single national hypertension program to be applied to all, and call for targeting responses to hypertension at the zone level and by sex, integrating hypertension into existing maternal, child, and chronic-disease health platforms, and making explicit the need to build a resilient pharmaceutical supply chain as a key element of macroeconomic policy, not an afterthought.

## Declarations

### Ethics Approval and Consent to Participate

The secondary data used for analysis in this study are de-identified and publicly available from the 2023–24 Nigeria Demographic and Health Survey (NDHS). The original survey team obtained informed consent from all interviewees, and the original NDHS protocol was approved by the National Health Research Ethics Committee (NHREC), Nigeria, and the ICF Institutional Review Board (IRB). This analysis relied on de-identified secondary data that is accessible via registered, authorized access to the DHS Program repository, so it was not reviewed for additional institutional review.

### Clinical Trial Registration

N/A

### Artificial Intelligence Statement

This work is not generated by a Generative Artificial Intelligence (G.A.I.) or large language model (L.L.M.) tool. The information provided is the authors’ own views and opinions.

## Acknowledgements

The authors are thankful to Ahead Labs, IIT Roorkee, for providing Stata 15 and the necessary skillset for this project. The authors also acknowledge Marwadi University for providing the resources that helped in the successful conduct of the research.

## Data Availability Statement

Data analyzed in this study are available from the DHS Program (https://dhsprogram.com) but are not publicly available because of restrictions to protect the respondents’ confidentiality. Access is only granted with registration and approval by the DHS Program, upon reasonable request at https://dhsprogram.com/data/available-datasets.cfm. The datasets used were the Individual Recode (women’s) file and the Men’s Recode file of the 2023–24 Nigeria Demographic and Health Survey (NDHS). We abstracted additional literature and information from publicly available clinical trial information and World Health Organization (WHO) reports.

## Large-Language Model (LLM)

N/A

